# Maliqua: A study within Malakit, a project on malaria and gold miners in French Guiana

**DOI:** 10.1101/2021.05.16.21257287

**Authors:** André-Anne Parent, Muriel Suzanne Galindo, Yann Lambert, Maylis Douine

## Abstract

Malaria is endemic in French Guiana, especially within the gold mining community working illegally. Gold miners travel to remote camps in the forest to carry out their activities, exposing themselves to the presumed contamination area. This paper presents the results of a qualitative case study of the Malakit project, a free distribution of self-diagnosis and self-treatment kits, along with appropriate training/information from health facilitators, at resting sites in Brazil and Suriname on the borders with French Guiana. This study documents how Malakit is part of the care trajectory of gold miners. The data was collected using three methods: 1) on-site observation; 2) semi-structured individual interviews (n=26); 3) semi-structured group interviews (n=2). The results inform us that Malakit responds to the need for treatment and facilitates access to care. Gold miners say they trust the facilitators and receive accurate explanations. The majority of participants find the kit easy to use and to carry and explanations given were sufficient, although some people needed to be reminded how to use it once in the forest. Results remind us that malaria among illegal gold miners in French Guiana is a question of social inequalities in health, where the interaction of the health, social, economic and political contexts of Brazil and French Guiana influence exposure to numerous risk factors. Thus, malaria intervention practices such as Malakit cannot be carried out without considering the complexity generated by social inequalities in health.

## CONTEXT

Malaria is the most widespread parasitic disease in the world, with 228 million cases worldwide in 2018 (World Health Organisation (WHO), 2019). While the majority are diagnosed in Africa, the South American continent is not to be neglected (Yan et al., 2020). In French Guiana, a French overseas territory in South America, malaria is endemic, especially within the gold mining community working illegally. Mostly coming from the poorest states of Brazil, miners are severely affected, with malaria prevalence ranging from 4% to 47% in this population (Douine, Mosnier, et al., 2018; Douine et al., 2016; Olapeju et al., 2020; Pommier de Santi et al., 2016). The literature reports that miners are reservoirs for malaria on the Guiana shield (Douine et al., 2020; Douine et al., 2016; Olapeju et al., 2020; Pommier de Santi et al., 2016) and while the region aims to eliminate malaria, the high prevalence of the disease in this hard-to-reach population, in conjunction with difficult access and incorrect use of antimalarial treatments, could favour the emergence of resistant parasites. However, the widespread and appropriate use of Artemisinin Combination Therapy (ACT), with the distribution of impregnated mosquito nets, could have an impact.

The French Guiana territory is largely covered by the Amazon rain forest and is located between the rivers bordering Brazil and Suriname, the Maroni and Oyapock rivers (Jolivet et al., 2012). Undocumented gold miners coming from Brazil travel these rivers by pirogues and the land on foot for many days at a time to get to the remote mining camps and carry out their activities in the forest, exposing themselves to the presumed contamination area (Douine et al., 2016; Douine, Sanna, et al., 2018). While doing so, they contravene with French regulations, conferring them an illegal status, and expose themselves to reprisals by the police authorities and the French army.

The issue of illegal workers in gold mining sites as a contributing factor for continued malaria outbreaks is largely discussed in local health authorities, but there is no national plan for malaria, since cases recorded in metropolitan France are only imported cases. Although the French Guiana Regional Health Agency has the authority to define public health strategies and recognizes the risk of antimalarial drugs resistance emergence, the required strategy and resources still remain to be defined.

Due to the geographical and regulatory issues in French Guiana, lack of knowledge about health care, poverty, and the legal status of gold miners, usual malaria control strategies cannot be implemented in this particular context. For instance, a strategy based on non-medical health workers, trained to diagnose and treat malaria in remote areas as implemented in Suriname is not allowed, because a non-health professional cannot provide diagnosis and treatment in French territory (Douine, Mosnier, et al., 2018).

Despite being a priority population in malaria elimination, there is scant literature on malaria-related behaviour among gold miners (Olapeju et al., 2020). Therefore, better knowledge to design tailored strategies targeting this specific population in the forest are required (Douine, Mosnier, et al., 2018; Douine, Sanna, et al., 2018). This article presents the results of a qualitative study conducted on an innovative community-based intervention tested within this population in French Guiana.

### THE INTERVENTION: THE MALAKIT PROJECT

In order to deal with the issues associated with malaria in the illegal mining community, a pilot intervention project called Malakit was implemented to evaluate a new malaria control strategy among gold miners working illegally in French Guiana. The working hypothesis of this project was: The free distribution of self-diagnosis and self-treatment kits, along with appropriate training/information from health facilitators, at resting sites in Brazil and Suriname, would be a strategy relevant to the specific context of French Guiana, and would address the identified need. The evaluation of the Malakit intervention was based on a series of outcomes measured by 1) a comparison of before/after cross-sectional estimates of self-medication behavior of gold miners (Orpal studies) and 2) a longitudinal monitoring of both safety and adherence to correct kit use (Douine, Sanna, et al., 2018; Galindo et al., 2021)(Malakit study^1^).

The main objective of Malakit was to increase the percentage of gold miners working illegally in French Guiana who take ACT entirely after a positive rapid diagnostic test. The secondary objectives were to reduce the prevalence of malaria among illegal gold miners in French Guiana and increase the percentage who: 1) know malaria, its causes, the symptoms and protection measures; 2) have a positive attitude towards recommended prevention measures, the use of adequate antimalarial treatment, and adherence to treatment; 3) report applying good preventive practices.

### THE INTERVENTION SITES

The Malakit project targets gold miners working illegally in French Guiana, a large majority of whom are Brazilian (+/−95%). The journey of these gold miners from Brazil to French Guiana passes either through the Oyapock River, between France and Brazil, or through the Maroni River, located between France and Suriname, or more rarely, by plane from Brazil to Suriname and then by car and/or pirogue to the river.

The areas through which the gold miners pass are therefore located by the two rivers, which also serve as rest, supply, and trading areas. On these sites, located on the Brazilian and Surinamese sides of the border rivers, the gold miners are not in an irregular situation and are therefore better able to take care of their health. For this reason, the distribution of the kits takes place on these sites. Although almost all gold miners are Brazilian, the political and social context differs according to the border under consideration:

#### In Suriname

Malakits are distributed in Albina, Antonio do Brinco and Paramaribo. Brazilian gold miners only need a valid passport to enter legally Suriname, but still find themselves in a foreign country with a different language (Galindo et al., 2021). The Surinamese Ministry of Health is very involved in the fight against malaria and the thousands of gold miners passing through Suriname represent a public health issue. This is why the Surinamese health authorities are invested in Malakit, with the scientific and operational support of the Foundation for Scientific Research in Suriname (SWOS).

#### In Brazil

Malakits are distributed in Oiapoque and Ilha Bella. The management of the Malaria Program in Brazil has ramifications at different state and municipality levels, which can complicate local collaborations with the Malakit project. The distribution of the kits in Brazil is carried out by a local association of development, accompaniment, animation and cooperation (DPAC Fronteira).The scientific counterpart in Brazil was the Foundation Oswaldo Cruz in Rio de Janeiro.

### METHODOLOGY

In order to improve the Malakit project, a qualitative study was undergone, using Yin’s case study methodology (Yin, 2018). This approach is most appropriate when the research question requires that attention be paid to the lived experience of actors; when the boundaries between the phenomenon and the context can be unclear; and when the goal is to describe and explain how everyday practices in specific places are connected to larger structures and processes. Furthermore, a case study as defined by Yin corresponds well to an interventional research approach.

This project documents how Malakit is part of the care trajectory of gold miners. It integrates with the broader Malakit study, offering a qualitative component to hear the opinion of gold miners and health facilitators on the use of Malakit. Within this project, the intervention is defined as the distribution of kits by health facilitators to gold miners working illegally in French Guiana in rest and supply areas outside the forest. When the kit is distributed, training on how to use the kit is offered by the facilitator and a first collection of quantitative data is carried out to ensure monitoring of the project. A phone app specifically developed for the project, usable without connection, includes a tutorial on the use of the kit and collection of the data of use.

The qualitative case study had the following objectives: 1) Understand the way gold miners perceive Malakit; 2) Identify the elements that are favourable and unfavourable to the use of Malakit; 3) Determine the contextual elements influencing the use of Malakit; 4) Identify what can be improved in the project. Data was collected in the same manner at both sites in Suriname (Antonio do Brinco) and Brazil (Oiapoque). The data was collected using three methods: 1) on-site observation; 2) semi-structured individual interviews; 3) semi-structured group interviews.

Two observation periods were carried out at each of the sites. Each of the observation period was carried out by the principal investigator, accompanied by a translator. The first period took place in April 2019. The objective was then to discover the context and places of distribution, to observe the distribution activities, to create links with the facilitators, and finally to inform the community about the qualitative component of the project. The second period took place in August 2019. The objective was to observe, carry out the interviews, and better understand the context of the interventions. During each period, notes were taken, interviews were recorded and transcribed and integrated into the dataset. The investigator spent time with the facilitators when she was not doing interviews and accompanied them to meetings with the gold miners.

Semi-directed interviews were conducted at each site in August 2019. The location of the interview was chosen by the participant and translated on-site from French to Brazilian Portuguese and vice versa. Using a method based on convenience samples, health facilitators made initial contact with gold miners who had already received a kit and who agreed to participate in the interview. Twenty interviews were conducted with both male and female gold miners. Twelve were interviewed in Oiapoque and eight in Antonio do Brinco, until data saturation. Six local actors were interviewed (three per site) based on their ability to influence the use of the kit or not. These people were hotel or canteen keepers or piroguiers transporting the gold miners and their equipment into the forest. Interviews lasted 30–40 minutes on average and each participant received compensation in the form of a 10-euro meal voucher.

Two group interviews were conducted at each site during working hours. Four health facilitators participated in Brazil and two in Suriname. In Brazil, the interviews were conducted in French with a translator, but in Suriname, they were conducted in English. Health facilitators either received a meal voucher or the investigator paid for the meal on their behalf, depending on the site. Interviews lasted an average of two hours and took place either at the Malakit office or in another remote and quiet location.

All interviews were recorded and transcribed when the principal investigator returned to the university in her country, except for one interview where the gold miner preferred that the interviewer take handwritten notes. A person of Brazilian origin was able to transcribe the interviews in Portuguese and French. The thematic analysis method of Mucchielli & Paillé (Mucchielli et Paillé, 2016) was applied using a semi-open descriptive coding grid based on ten general codes (Miles, Huberman et Saldana, 2014).

## RESULTS

The results are presented according to a summary of the information referenced for each code in relation to the research objectives. It is important to mention that overall, the participants say that Malakit is a success and that it is important to maintain the project.

### 1. The context and history of malaria among gold miners

Only two of the twenty gold miners interviewed said they had never contracted malaria. A large majority of them have contracted it several times in their lives. Before Malakit, the gold miners had little choice but to leave the forest to be treated in a health centre (a journey that can be very long, sometimes many days on foot and/or by boat) or to be treated with medication bought illegally on gold mining sites in the forest, in particular Artecom®, which contains artemisinine derivatives (Douine, Lazrek, et al., 2018). Gold miners rarely used the entire treatment because of the high cost: either they save a few tablets for the next crisis or they share it with another person.

All the participants identified malaria as the most important health problem for gold miners. This is a major constraint as many participants mention that health is the most important thing they have because a good state of health allows them to work. Hence, malaria has an impact not only on their health, but also on their ability to work, which in turn impacts their economic situation, as they are paid according to the gold collected. If the chosen treatment is not effective in treating the problem, or if the symptoms of the disease or the side effects of the drugs make them unable to work for several days, they have to deal with consequences in terms of income. Health facilitators report, however, that gold miners sometimes ignore or confuse the symptoms of malaria with other tropical diseases. The self-diagnostic test and the paracetamol tablets (to be taken as an alternative in case of a negative test for malaria) included in the kit are therefore very important.

> “Then I started to feel very cold, I felt very bad. I used it on myself, but the test was negative” (Gold miner, P14).

Buying medicine in the forest can be very expensive since it is done outside the regulated market. Distributed free of charge and considered effective, Malakit is thus seen as the best solution against malaria. Some participants even attribute to it the observed decrease of malaria cases in the region.

### 2. The perception of the Malakit project and its health facilitators

Word of mouth among the gold miners, the posters hung in the community, the large banner in front of the office in Oiapoque and the presence of a health facilitator in the community, often considered as a good acquaintance or even a friend, are the main sources of dissemination of information about Malakit. The bond of trust with the facilitators is essential. The gold miners say they trust the health facilitators, despite some fears related to their own administrative status (illegality on French territory), which quickly dissipates after the first contact.

> “I saw a Malakit poster. Friends also told me about it” (Gold miner P01).
>
> “It is thanks to [the health facilitator]. I saw several friends using this kit” (Gold miner P05).

The contact with health facilitators is generally positive. Participants report that they are treated well, receive accurate explanations, and that they appreciate the tutorial and other visual aids used to explain the test and treatment. Some facilitators use the image of friendship to describe the bond they create with the gold miners. This shared bond is confirmed by the comments of gold miners who express their satisfaction with all the facilitators and the importance of the links established with them. Facilitators insist on the importance of explaining the use of the kit simply and quickly. Communication, openness and adaptation to the realities of the people encountered seem to be central to the health facilitators’ approach. Some of them, particularly the ones who have practised gold mining before, take a great deal of pride in prevention and knowledge dissemination.

> “It is at work that we develop a relationship of trust and friendship […]. For me, it’s important, because I gain the confidence of the gold miners […] To be able to bring knowledge, to exchange knowledge with a [social] class that is discriminated against, the class of gold miners. For me, it is a pleasure to work at [distribution site]” (Health facilitator M01).

In general, the key actors we met do not frequent the Malakit premises and the majority do not go and get the kit for themselves because they are at low risk of contracting malaria. However, they all mentioned having a very positive attitude towards the project. They define themselves as “intermediaries” for the facilitators. Since some of them have worked or still sometimes work at the gold mining sites, they are important allies in learning about the reality and needs of the gold miners, the routes taken, and how to use the kit. These key actors could therefore be more involved in organizing and advertising the distribution of the kits in a collective effort to educate, particularly about the risks of sharing the medication.

Despite the general enthusiasm for the project, however, some participants mentioned that the training is easy to understand when the health facilitator explains it, but when the gold miners use it in the forest later on, they forget the instructions and have difficulty using the kit, despite the video and other material given. Many said that the spirit of solidarity within the forest therefore becomes essential so that the less educated can use the kit. It is also important to mention that the application is more often installed on cell phones in Suriname (almost systematically) than in Brazil. However, it was not possible to explain this difference.

Moreover, while some gold miners might at first sight fear a certain complicity between the French judicial authorities and the Malakit project, they generally developed trust after a certain time. According to the facilitators, the perception of the project has evolved over time. In the beginning, people were distrustful, but less and less so with time, and today, the gold miners who fear the project are said to be marginal. Gold miners still fear the police and the army or anything that might prevent them from working, which makes them constantly hypervigilant, but health facilitators have generally managed to cope with this constraint. Facilitators, however, mention differences between Oiapoque and Ilha Bella or Antonio do Brinco. For the first site, located in town, people come to the Malakit premises, while for the second and third, it is rather the facilitators who go to the gold miners, or the inhabitants of the village who bring them to the facilitators. This difference between the sites located in the city and those outside suggests that the notion of proximity plays an important role in the creation of links and, by ricochet, in the distribution of the kits. The distribution of the kits by the facilitators in the community could thus be more effective than their presence in an office.

> “No worry, because I’ve known X (Health facilitator) since I was very young” (Gold miner P15).

> “What I can say now is that the gold miners are very accepting this project. Very well. They are very interested in the project … […] we are seen as friends, mentors, psychologists. A person who is there just to help them […] It’s an interesting and very important project for them” (Health facilitator M01).

### 3. Elements favourable and unfavourable to the use of the kit

The majority of participants find the kit easy to use and to carry in their bags. Many participants returned for a second kit, satisfied with their first experience. Some said they used the first kit when they were sick, while others gave it to another gold miner who was sick. In general, the explanations given by the facilitators when they received the kit were sufficient. However, some people need to be reminded how to use it, but there is always someone at the gold panning site who received the kit and knows how. The only fear mentioned in its use is when it comes to “pricking the finger and putting it in the hole” so that the blood can be deposited on the diagnostic test strip.

> “Everything was in the video […], but I still watched because I was afraid of making a mistake […] it’s very easy” (Gold miner P13).

On the issue of medication, several participants mentioned that gold miners share medication at gold mining sites. When asked, participants admit that this practice exists, but they do not seem to be aware of the impacts of such a practice. In fact, the majority of the miners interviewed said that when they return to the site in the forest, they can share—or give—their kit to someone in need and that solidarity in the face of illness is what matters most to them. People who donate their kit will often do the test themselves on the person with symptoms and will explain the project and how to obtain a kit. A minority of participants mention having seen kits sold in whole or in part, while others say they have never seen this phenomenon. Many report that their equipment was burned by the military during an intervention at the sites or left behind when they fled. The question of access to the kit is raised in a tangible way, particularly for people who do not frequent the resting sites and who remain on the gold mining sites in the forest. While it may be thought that there is some form of “traffic”, the interviews highlight the solidarity between the gold miners when they are in need. However, some key actors are concerned that the kit may be resold and consider that those in charge should distribute more kits to deal with this phenomenon. In addition, sharing of kits, through donation or sale, exists and it is important to take this into account because Malakit managers and facilitators do not control the “distribution chain”. Health authorities, decision-makers and donors should therefore be made more aware of the contextual elements related to survival in the forest.

> “Yes, I did […] I took two doses and gave two doses to the other person […] Everyone knows it’s hard to get (the kit). So when one person is very sick, we share our medicine” (Gold miner P01).

It is easy to say that the perception of the Malakit project by the gold miners is very positive and its effectiveness against malaria is recognized in the community. They said the medication works when they are sick, and they like the fact that the kit is free and relatively easy to access. All the participants mentioned that they talk about the kit extensively to their entourage and encourage the gold miners to go and get it when possible.

> “We talk about it all the time, especially when we’re in the forest. We talk to friends about coming to get the malakit […] Everyone speaks well about it. Everyone comes to pick them up before going into the forest” (Gold miner P15).

However, according to the people interviewed, knowledge of the Malakit project seems to vary from one site to another. Some gold miners claim that everyone knows it, others are more nuanced. In fact, some miners know it exists and would like to get it but are in a hurry when they are on the resting sites in transit, or unable to come and get it. Training time (30-40 minutes) also comes up as a discouraging factor. However, the main obstacles seem to be access to the resting sites for many, as well as the dangers associated with police and army inside the forest or on the roads leading to them. Many participants mentioned the difficulty for some miners to obtain the kit because they never leave the forest. They suggest a wider distribution, particularly at the illegal gold mining sites in the forest. They think that the facilitators could do the distribution at these sites or that the miners themselves could bring the kits and distribute them. Participants generally agreed on this issue and seem unaware of the difficulties associated with their proposal.

Key actors are important when considering access because they share information about the Malakit project, in addition to sometimes showing gold miners how to use the kit. According to some, there are still many people at gold mining sites who do not know about it. In their view, most gold miners would like to take it with them to the gold mining site just in case, and they do not hesitate to refer them to the facilitators. While the perception may have been negative at the beginning of the project’s deployment due to fears of a possible police presence, this is no longer the case, thanks, in particular, to the key actors who encouraged the gold miners to use it. The link between the key actors and the health facilitators is therefore essential and must be maintained.

### 4. The context, living conditions, and other health problems

Most participants mentioned experiencing other health problems related to either their gold mining activities or to the living conditions of gold miners in general. The main health problems mentioned were HIV, hepatitis, kidney and liver problems, general physical pain, back pain, leprosy, leishmaniasis, dengue fever and anemia. Several people mentioned a lack of information about the diseases and confusion about the symptoms related to the different diseases. Many also mention problems related to lack of water or lack of drinking water because they are regularly forced to drink water and wash in rivers where they pan for gold or small creeks near camps and toilets.

> “The first thing they ask is if we have the test for HIV… And if we have tests for leishmaniasis” (Health facilitator, M01).

> “The person who catches malaria and doesn’t pay attention, they will end up sick of something else… I see that there are many sexually transmitted diseases… Because condoms are not easily distributed among them. Especially here. We don’t have resources. The health is terrible. The hospital takes care of it just if it is a case of great danger” (Key actor, A2).

> “There are a lot of diseases that we see at the gold panning site and we don’t even know what they are […] (Gold miner P17).

Although the data collected on women’s health problems is not consistent, it appears that women’s situations are different from men’s situations. Many mentioned having problems specifically related to menstruation or lack of hygiene, and some spoke of women engaged in sex work at gold mining sites and in resting zones, mentioning that HIV and other sexually transmitted diseases are difficult to prevent because they encounter men who refuse to use condoms. In addition, some women reported being abused or assaulted by men, sometimes police or soldiers, while fleeing in the forest.

Above all, participants mention that their main needs are money and work. However, several other themes emerged from the data, such as the need for social recognition and the fear of the police destroying or burning their goods. On that subject, health facilitators report that the discrimination experienced by gold miners, the inability to have their rights recognized, and the lack of institutionalized organizations lead them to be considered as “zombies” for society. The gold miners suffer psychologically, physically, and socially because of the legal, economic, and social exclusion they experience. Moreover, facilitators consider gold miners to be vulnerable and easily exploited, needing legal, political, and social recognition. The key actors met recall that police interventions cause instability, and create people on the run, who live in constant fear and anxiety. The lack of employment and the disparity between living conditions and gold mining in French Guiana and Brazil are said to be the main causes of these inequalities. Finally, it should be reminded that not everyone in the forest has the same status and that there are armed militias looting in the forest, putting the gold miners at risk. Unfortunately, for a large part of the population in French Guiana, the miners are not considered the victims of this phenomenon but rather the cause. It is therefore very difficult for the gold miners to “get in” and “out” of the forest and to seek treatment for their health problems. Many remain ill, without leaving the area, only to come out again in a situation of imminent death.

> “The gold miners are not well seen. And their rights are being taken away … their rights to health, their rights to education, the right to leisure. We who work as health facilitators have become the psychologists of the gold miners” (Mediator M01).

> “The police don’t let them go to work and they have to live on the run. It’s a difficult situation. They come and they break and they burn everything […] They have no job opportunities, they have no profession and many of them are forced to work on the other side, even illegally” (Key actor A01).

### 5. Malakit elements that can be improved

When participants were asked what they would change in the Malakit project, the vast majority responded: increase distribution! Whether on gold mining sites in the forest or elsewhere, all considered that Malakit should continue and expand its distribution network. The project addresses the gold miners’ need for treatment, facilitates access to care, and saves lives overall.

Some people made suggestions, ranging from adding various drugs, water treatment tablets, and iron supplements to existing kits, to develop kits for other diseases, including leishmaniasis. Some also mentioned that it would be interesting to have health facilitators who could address a variety of questions, whether for health or other social problems. In fact, some facilitators from the Malakit project are already helping the gold miners with a wide range of problems, the most important being the loss of identity papers once the army has burned their property in the forest, many gold miners being illiterate, and needing help dealing with institutions.

Those interviewed did not express an opinion on the deployment of Malakit to other regions. Many deplored the fact that there were a limited number of gold miners in Oiapoque when we visited during the summer of 2019 due to the police presence upstream, but did not propose new distribution sites. Since the presence of gold miners in a region is dependent on the possibilities of gold mining and police constraints, it seems that the people we met have little power to influence this reality. However, the key actors in Oiapoque, who are dependent on trade with gold miners, expressed fears about their ability to remain in the region and continue their commercial activities as owners of hostels and pirogues.

## DISCUSSION

Reading the results of this project, one cannot help but recall what the WHO Commission on Social Determinants of Health said a decade ago, highlighting the fact that inequalities kill on a large scale (2009). The World Malaria Report (WHO, 2019), for its part, informs us that “malaria is a disease that is increasingly emblematic of poverty and inequality, characterized by higher morbidity prevalence in the most vulnerable populations”. Thus, malaria among gold miners working illegally in French Guiana is truly a question of social inequalities in health, where the interaction of the health, social, economic, and political contexts of Brazil and French Guiana influence exposure to numerous risk factors.

As a reminder, social inequalities are the result of “an unequal distribution of resources, produced by society and giving rise to a sense of injustice” (Moulin, 2014). Social inequalities in health, on the other hand, refer to differences in health within a population caused by the conditions in which individuals live (WHO, 2009). These gaps, which are found between men and women, between socio-economic groups and between territories, are socially constructed, fundamentally unjust and avoidable; they are primarily a matter of social justice (De Koninck, 2008; Parent et Bourque, 2016).

As the role of the social health determinants in the production of social inequalities in health is widely reported in literature (Baum, 2008; Gore et Kothari, 2012; Marmot, 2009; Potvin, 2010), it is recognized that social inequalities in health within and between societies exist and are perpetuated by supportive government policies (Navarro, 2009). Social inequalities in health would thus be the result of a double burden: the socio-economically disadvantaged are both more exposed to stressful living conditions and less endowed with protective resources. The social inequalities present in a society are thus transposed into inequalities in living conditions and, finally, create social inequalities in health.

Income inequalities among the Brazilian population are leading individuals to choose illegal gold mining in French Guiana to provide for themselves and their families. Thus, the living and working conditions of gold miners predispose them to malaria and other diseases. In other words, the determinants of malaria are essentially social in nature and are fuelled by poverty. Among the determinants observed or named during data collection are low education, unemployment, migratory status and gold mining regulations in French Guiana, poor housing on gold mining and resting sites, working conditions, water quality and fecal peril, and gender inequality. Malaria intervention practices cannot therefore be carried out without considering the interaction between these health determinants and the complexity generated by social inequalities in health.

Despite the efforts already made in the design of Malakit, the issue of health literacy remains and could be given more consideration because low health literacy is a barrier to the self-management of health problems (Lloyd, Ammary, Epstein, Johnson et Rhee, 2006). Defined as the cognitive and social skills that determine an individual’s willingness and ability to identify, understand and use information to promote and maintain good health (Nutbeam, 2000), health literacy involves taking into consideration the contexts, both environmental and individual, in which a project takes place. Thus, as the WHO (WHO, 1998) points out, health literacy means more than the ability to read the documents provided. By improving access to health information and, more importantly, the ability to use it effectively, health literacy is essential to empowerment (Mosnier et al., 2020).

Health literacy is itself dependent on broader levels of literacy. Low literacy levels can directly affect people’s health by limiting personal, social, and cultural development and by impeding the development of health literacy (WHO, 1998). Many people have pointed out gold miners’ low level of education and their need for health support. In addition, several gold miners expressed fears related to the use of the kit, mainly about the self-diagnostic test. In order to further integrate this notion into the project, it might be relevant to train the health facilitators to better develop the skills necessary to use the kit. For example, and even though this is already done during the training, the finger prick that many people are concerned about could be further demystified and practised to a greater extent with a facilitator.

## RECOMMENDATIONS AND CONCLUSION

In its current form, Malakit undoubtedly contributes to the reduction of social inequalities in health because it combines an action of self-diagnosis and self-treatment of malaria with an outreach intervention among a marginalized population. While this project was able to document the intervention and the perception of Malakit, certain reflections and recommendations emerged. You will find them in the next paragraphs.

The community intervention offered by facilitators is essential. This intervention is influenced by the time spent in the area, the ability of facilitators to create bonds with the gold miners and key community actors, the tools and concepts of popular education available to them, their openness and knowledge of the gold miners’ reality, and finally, the presence of local partners from the health network, who may be favourable or unfavourable to the project. As the strategies for disseminating the kits differ depending on whether they are inside or outside of the cities, it is relevant to reflect on the contexts, their possibilities, and constraints in a specific way. Information on the risks of sharing medication could be better disseminated, as sharing exists. Since sharing can be interpreted as a solution developed by gold miners in the face of difficulties in accessing care and treatment and highlights their solidarity in the face of adversity, it is important to understand the context. The police presence on the routes taken by the gold miners contravenes their ability to access treatment and take care of their health.

In order to reach more gold miners, a flexible system of distribution could be envisaged in order to quickly adjust to their movements on the territory. Thus, some sites would be permanently invested, while other sites would be temporary. Interventions would have to vary according to the type of site. Peer interventions at gold mining sites could also be relevant. These peers could play an important support role for those who have more difficulty using the kit. They could also disseminate prevention messages or share relevant information on different determinants of health. Recruited among the gold miners, these people could be better recognized and put to work within the project. Even if the majority of participants found the kit easy to transport to the gold mining site and simple to use, strategies that support health literacy could be improved for the use by illiterate people and promote self-management.

Gold miners have many other problems than malaria for which they have difficulty obtaining care. More partnerships with local health organizations or associations would be relevant, such as organizations that carry out prevention and testing for HIV and other STIs. In addition, access to safe drinking water is a major issue. Water treatment strategies, such as the insertion of water purification tablets in the kits, could be evaluated and developed within the Malakit project. Several people also mentioned that strategies to treat anemia should be considered as there are close links between malaria and anemia. Finally, and not the least, the situation of women at gold mining and resting sites should be further studied to better understand their specific risk factors.

In conclusion, we would like to remind that all participants identified malaria as the most important health problem for gold miners. This not only has an impact on their health, but also on their ability to work and their economic situation. The perception of the Malakit project by the gold miners is very good and its effectiveness against malaria is recognized in the gold mining community. Although this project is not a miracle model, we can say that it is a promising strategy for improving the health of gold miners. The quantitative results of this study, which are very promising, will be published shortly.

This being said, the main needs of gold miners are financial. In order to meet these needs, they face multiple obstacles and put their lives at risk in a variety of ways. The exclusion mechanisms resulting from this situation are deeply rooted in local, regional, national, but also cross-border economic and political systems. The reproduction of social inequalities in health cannot be resolved without a real willingness on the part of health and political decision makers. The promoters of Malakit must also deal with this reality and the constraints it generates. Nevertheless, Malakit is an innovative and highly promising intervention to increase access to health services, diagnosis, and treatment of malaria, as proposed by the “World Malaria Report 2019” (WHO, 2019). If the project is already being pursued in Suriname by the National Malaria Control Program, it still needs to be accepted and financed by the French and Brazilian authorities for its sustainability.

## Data Availability

Permission for access to data can be obtained through corresponding author

## Footnotes

1 https://www.malakit-project.org/evaluation/

